# Impact of Perioperative Dexamethasone in the Context of Neurosurgical Brain Metastasis Resection

**DOI:** 10.1101/2023.01.23.23284887

**Authors:** David Wasilewski, Jan Bukatz, Ricarda Peukert, Zoe Shaked, Paul Poeser, Anna-Gila Karbe, Anna-Trelinska-Finger, Claudius Jelgersma, Anton Früh, Matthias Raspe, Helena Radbruch, David Capper, Max Schlaak, Peter Thuss-Patience, Philip Bischoff, David Horst, Marcel Krenzke, Ran Xu, Felix Ehret, David Kaul, Martin Misch, Lars Bullinger, Nikolaj Frost, Peter Vajkoczy, Julia Onken

## Abstract

**Background:** Patients with brain metastases that undergo brain metastasis resection regularly receive perioperative dexamethasone. We sought to evaluate whether perioperative dexamethasone in brain metastases is linked to survival.

**Methods:** Retrospective data on perioperative dexamethasone dosage in resected brain metastasis patients at three hospital sites of the Charité from 2010-2022 were collected. Cut-off values for cumulative perioperative dexamethasone dose as a continuous predictor variable for survival were determined using maximally selected rank statistics. Patients were dichotomized based on determined cut-offs of cumulative dexamethasone (pre-operative: < 40 mg vs ≥40 mg; post-operative: < 180 mg vs ≥ 180 mg) and pre- and postoperative: < 281 mg vs ≥ 281 mg). Medical records included baseline demographic, radiological, histopathological and treatment-related characteristics. Based on cut-off values for dexamethasone downstream statistical analyses included Kaplan-Meier, Cox proportional hazards regression for overall survival with adjustment for potential confounders including age, gender, Karnofsky performance status and presence of extracranial metastasis via propensity score matching.

**Results:** 539 patients were included. Median follow-up time was 58,97 months. After adjusting for age, gender, Karnofsky performance status and presence of extracranial metastasis patients with higher cumulative perioperative dexamethasone (≥281 mg) showed shorter survival (HR: 1.47 (1.20-1.80, p<0.001) as compared to patients with lower cumulative doses (<281 mg). This effect remained significant after correction for patients that died within 2 months after resection and for patients with KPS below 50% and, independently from this approach performing propensity score matched-based analysis of the total cohort of patients, respectively.

**Conclusion:** Cumulative perioperative dexamethasone is associated with decreased survival in the context of brain metastasis resection. Strict dosage, down taper or methods reducing corticosteroid dependency should be regularly evaluated in clinical practice in patients with brain metastases.

## BACKGROUND

Up to 50% of cancer patients develop brain metastasis within the course of disease^1,2^. Brain metastases are in general associated with a bad prognosis, however a subset of these patients with good clinical performance or good Karnofsky performance status (KPS), surgically accessible and/or symptomatic brain metastases are regularly undergoing extensive treatment including neurosurgical brain metastasis resection followed by radiotherapy (RTx)^3-5^. Apart from advances in local therapy (e.g. stereotactic radiosurgery (SRS)), systemic treatment approaches have experienced some major changes. Adjuvant therapies have shifted from post-operative RTx and chemotherapy (CTx) to post-operative RTx with targeted therapies including small molecule inhibitors or checkpoint inhibition (CPI)^3-7^. Similar to glioblastoma treatment, patients with symptomatic brain metastases that undergo neurosurgical treatment with craniotomy and microsurgical brain metastasectomy regularly receive pre- and postoperative dexamethasone to treat perifocal edema to alleviate neurological symptoms^8,9^. Yet, dexamethasone use may be associated with significant adverse effects and immunosuppression^10,11^. In addition, there is ample evidence from the field of glioblastoma to suggest that pre- and postoperative dexamethasone is associated with worse overall survival (OS) as recently shown in several retrospective studies^12-14^. No larger randomized-controlled prospective trials have been conducted on this matter up to now. This holds also true for patients with brain metastases. Importantly, systematic studies evaluating potential adverse effects of dexamethasone including its impact on OS, other complications such as negative impact of perioperative dexamethasone on post-operative treatments including CPI could provide important information and generate more evidence for decision-making with respect to dexamethasone dosage regimens. Given the paucity of data, we aimed to investigate whether perioperative dexamethasone (i.e. administered in the pre-operative period, post-operative period and in both periods) adversely affects clinical outcome in patients undergoing brain metastasectomy. Real-world data on perioperative dexamethasone dosage may aid in further evaluating dexamethasone dosage and initiating randomized-controlled studies to find an optimal dosage regimen for these patients. This retrospective, comparative effectiveness study describes detailed perioperative dexamethasone dosage in a large cohort of patients who underwent brain metastasectomy (n=539). Further the study provides descriptive information on demographics, radiological and histological features as well as treatment-related features after brain metastasis resection. To find optimal cut-off values or cut-points based on different settings or time windows for dexamethasone dosing in the context of metastasectomy, we used a well-known cut-off estimation model by Hothorn and Lausen called the maximally selected rank statistic. Using this model, we examined the association between cumulative preoperative, postoperative, and total dexamethasone dose and clinical outcome in conjunction with a downstream descriptive and inferential statistics with overall survival as the primary outcome ^15^.

## METHODS

### 1.1 Patient cohort and study variables

For this retrospective study all patients with neurosurgically resected brain metastases treated at all three sites of Charité-Universitätsmedizin Berlin in the period from January 2010 to December 2022 were included. Data censoring was 1^st^ of December 2022. Patient data were identified as previously described using an institutional database (SAP, Walldorf, Germany) as well as the Charité Comprehensive Cancer Center (CCCC) Registry^16^. Patients with a histopathological confirmation of an intracerebral manifestation of non-small cell lung cancer (NSCLC), breast cancer, melanoma, renal cell carcinoma (RCC), colorectal cancer (CRC), esophageal cancer or gastric cancer or unknown primary tumor were included. This study is in accordance with the ethical standards outlined in the Declaration of Helsinki and was approved by the research ethic board at the Charité (EA1/399/20)^16^. Exclusion criteria are displayed in the consort diagram (**Figure 1**). Exposure to perioperative dexamethasone was defined as follows: pre-operative dexamethasone included cumulative dexamethasone dosage in mg from day thirteen before the day of operation (−13d) till the day of operation (d0), post-operative dexamethasone included cumulative dexamethasone dosage in mg from day one after the day of operation (1d) till day thirteen after day of operation (d13), and pre- and postoperative dexamethasone hence was defined as cumulative dexamethasone dosage in mg from day thirteen before the day of operation (−13d) till day thirteen after day of operation (d13) (**Figure 1**). OS represented the primary outcome of this study and was defined as time from brain metastasis resection until death from any cause. Baseline was defined as day first brain metastasis resection in each patient’s history; baseline characteristics were selected according to previous retrospective studies ^6,7,15^. KPS was assessed after first brain metastasis resection and dichotomized in good (≥70%) and bad (<70%). Presence of underlying other diseases was defined as presence of cardiovascular diseases, chronic lung, renal or liver diseases and was dichotomized in either not present or present. Radiological baseline characteristics included anatomical localization of resected brain metastases, number of brain metastases at baseline, presence of extracranial metastases at baseline and were based on reports from board-certified radiologists (**eTable 1a-b, eTable 2 a-b in the online supplements**). Tumor volumes and associated edema volumes were quantified using a semi-automated 3D rendering algorithm in iPlannet (Brainlab, Munich, Germany) using the SmartBrush tool (T1-weighted images for tumor and FLAIR-images for edema measurements). Only the resected lesion was quantified; in case of multiple lesions no addition of tumor or edema volumes was performed. Treatment-related features included the total number of brain metastasis resections, resection of primary tumor mass, administered systemic treatment modalities before brain metastasis resection and adjuvant therapy after brain metastasis resection. Information on systemic treatment and radiotherapy before or after first brain metastasis resection at our institution was retrieved from our database and patient records similarly to our previous study^16^ (**eTable 1a-d in the online supplements**). Follow-up data were retrieved from the registry from the Charité Comprehensive Cancer Center (CCCC) and obtained until December 1^st^ 2022. In NSCLC driver mutational status or included driver mutational status of epidermal growth factor receptor (EGFR), anaplastic lymphoma kinase (ALK) or c-ros oncogene (ROS1) translocations were regularly assessed after 2015. In melanoma v-raf murine sarcoma viral oncogene homolog B1 (BRAF) from 2014 onwards and human epidermal growth factor receptor 2 (Her2) status in case of breast cancer; thyroid transcription factor-1 (TTF1) status was regularly documented, whereas progesterone receptor (PR) and estrogen receptor (ER) status were documented in case breast cancer brain metastasis was suspected. Further histopathological information retrieved from patient records included Ki67 index and programmed death ligand 1 (PD-L1) tumor proportion score (TPS) of resected brain metastasis tissue (**eTable 1a-d**). Institutional pathological review was mandatory, all resected specimens were reviewed by board-certified neuropathologists for diagnosis. Chi-square test of independence was used to analyze the frequency table for categorical variables. Non-parametric Wilcoxon rank sum test was used for comparing two means not normally distributed data of this dataset; when expected count was below 5 Fisher’s exact test was used.

**Figure 1:**
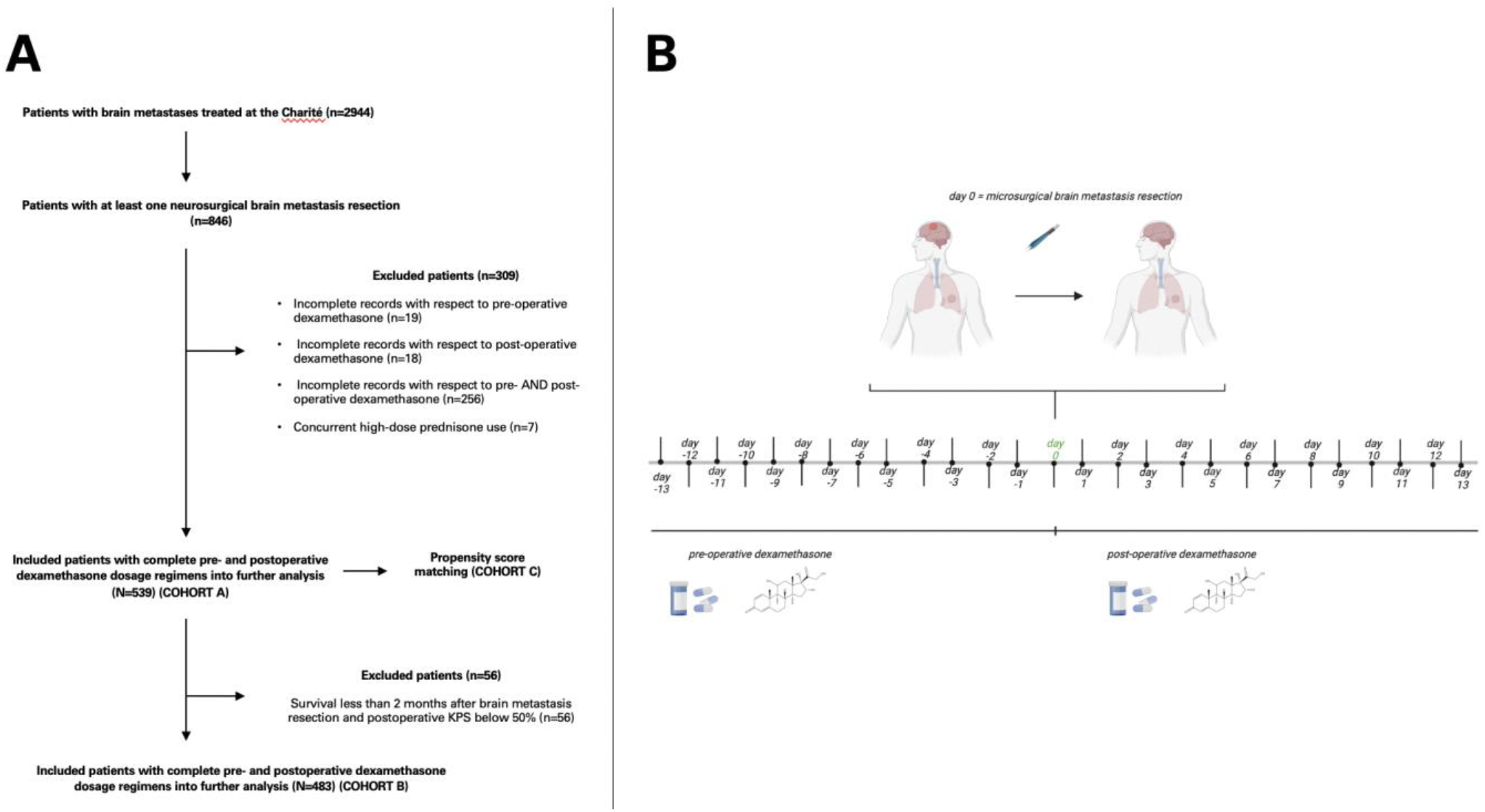
Flow-chart of exclusion and inclusion criteria and overview of the resulting groups of patients of patients with neurosurgically resected brain metastases in this study (COHORTS A, B and C) (**A**) graphical abstract of the study depicting pre- and postoperative dexamethasone dosing from day -13 to day +13 in relation to the date of operation (**B**).

### 1.2 Statistical analysis

We used R studio (Version 2021.09.2, R Foundation for Statistical Computing, Inc, Boston, USA) to compute descriptive and inferential statistics as in our previous study^15^. The *gtsummary* package was used to describe tabular data of our patient cohort, including categorical and numerical variables (https://cran.r-project.org/web/packages/gtsummary/index.html). Median OS was estimated by means of Kaplan-Meier analysis with confidence interval bands being displayed in the respective figures: Plotting was performed using the *survival* (https://cran.r-project.org/web/packages/survival/index.html)and *survminer* package (https://cran.r-project.org/web/packages/survminer/index.html). The prognostic value of each variable was tested using the log-rank estimator. Univariable and multivariable Cox regression modeling served to assess the effect of one or the simultaneous effects of multiple clinical variables on OS and was done using the *survminer* package. Cut-off values for cumulative perioperative dexamethasone dose as a continuous predictor variable for survival were evaluated and determined via an algorithm using maximally selected rank statistics implemented in the *maxstat* package (https://cran.r-project.org/web/packages/maxstat/index.html). Patient that were included in this study includes the total cohort “all” (N=539), referred to as COHORT A and a subcohort after excluding patients with less than 2 months OS and patients with KPS less than 50% named “2months and KPS<50excl”, which was termed as COHORT B. The effect of perioperative Dexamethasone on survival was also reevaluated via 1:1 (nearest-neighbor) propensity score matching as described in our previous work^15^. Propensity score matching involved the following baseline covariates: age, gender, KPS (dichotomized into <70% or “bad” and ≥70% or “good”) and presence of extracranial metastases. The matched cohort is henceforth referred to as COHORT C. Kaplan-Meier analyses were performed after applying different cutpoints and dichotomizing patients into subgroups according to optimal cut-point value for pre-operative dexamethasone, postoperative dexamethasone and pre- and postoperative (total) dexamethasone. Further R packages used, included common analytical packages such as *dplyr, tidyverse*. We conducted univariable and multivariable analysis for both datasets (COHORT A, B and C) for OS as a main endpoint adjusting for potential confounders such as age, gender, (KPS) and presence of extracranial metastasis was performed. Data collection was done with Microsoft Excel (Version 14.3.9, Microsoft Inc., Redmond, USA). Graphpad Prims (Version 9, Graphpad Software, Inc., San Diego, USA) was used for plotting Suppmentary figure 2b. A p-value of < .05 was considered significant with p-values being two-sided. R code and raw data will be made available at github upon request.

## 2 RESULTS

### 2.1 Baseline characteristics

Characteristics of patients with cancer brain metastases that met the inclusion criteria COHORT A (n=539) are shown in the supplementary material section (**Figure 1, eTable 1a-d in the online supplements**). There were 260 (48%) male and 279 (52%) female individuals in the total cohort of patients. Median age was 64 years (55-72, IQR). Most common symptoms associated with brain metastases were sensory-motor symptoms (21%), vertigo or dyscoordination (20%) and headache (12%). Sixty-two percent of patients had a good postoperative KPS (≥70%), whereas 38% of patients had a bad KPS (<70%). Predominant histopathology included brain metastases due to NSCLC, 93 (17%) were due to breast cancer, 91 (17%) were due to melanoma and 33 (6,1%) were due to SCLC. Renal cell cancer accounted for 4,8%. Two hundred eighty-nine (45%) patients were pre-treated with systemic anti-tumor therapy before first brain metastasis resection (baseline), and 238 patients (55%) were treatment naïve. Other radiological, histopathological, and treatment-related characteristics of this collective are summarized in supplementary materials (**eTable 1a-d**). Median follow-up time (IQR) was 58.97 months (31.97-89.17 months). Cumulated median OS of the COHORT A (n=539) was 13.3 month (95% CI: 12.1 – 15.2) and 11.4 (95% CI: 10.2 – 13.2) (p=0.049) for the COHORT B (**eFigure 1, eTable 2a-d in the online supplements).**

### 2.2 Dexamethasone dosage regimens and association of dexamethasone with overall survival

The median dexamethasone dose was 80 mg for the pre-operative period, 96 mg for the postoperative period and 190 mg for the total pre- and post-operative period for the COHORT A and 80 mg for the pre-operative period, 84 mg for the postoperative period and 187 mg for the total pre- and postoperative period for the COHORT B, respectively (**Table 1a, 1b**). The cumulative total dexamethasone dose prescribed during the pre-operative and post-operative period did not correlate with age, KPS or histopathological markers such as Ki67 or PD-L1 TPS, tumor volume or edema volume in COHORT A (**eFigure 2a, b in the online supplements**). 38 patients did not receive dexamethasone at all. Interestingly, within COHORT A and COHORT B no difference in OS was observed between patients that did not receive perioperative dexamethasone (−DEXA) vs. those patients that did receive perioperative dexamethasone (+DEXA) (**eFigure 3a, b in the online supplements**). Similarly, we did not observe any differences in OS within both cohorts when a cut-off of 8 mg dexamethasone daily was used to dichotomize patients during the pre-operative and post-operative period (**eFigure 3c, d in the online supplements**).

**Table 1a:** Comparison of the two patient groups of interest before propensity score matching within the whole patient cohort (“all”) (N=539). The optimal cutpoint value of 281 mg for perioperative dexamethasone was used to dichotomize patients. ^2^Interquartile range (IQR).

| Characteristic | Treatment |  |  | p-value<sup>2</sup> |
| --- | --- | --- | --- | --- |
|  | Overall, N = 539<sup>1</sup> | <281 mg, N = 402 | ≥281 mg, N = 137 |  |
| **age (years), Median (IQR)** | 64 (55 – 72) | 64 (55 – 72) | 65 (56 – 71) | 0.74 |
| **gender, n (%)** |  |  |  | 0.99 |
| female | 279 (52) | 208 (52) | 71 (52) |  |
| male | 260 (48) | 194 (48) | 66 (48) |  |
| **pre-operative dexamethasone (mg), Median (IQR)** | 80 (26 – 144) | 59 (24 – 115) | 216 (144 – 336) | <0.001 |
| **post-operative dexamethasone (mg), Median (IQR)** | 96 (62 – 132) | 84 (60 – 110) | 156 (108 – 230) | <0.001 |
| **peri-operative dexamethasone (mg), Median (IQR)** | 190 (132 – 283) | 160 (114 – 204) | 373 (320 – 438) | <0.001 |
| **KPS_group, n (%)** |  |  |  | 0.43 |
| ≥70% | 326 (62) | 246 (63) | 80 (59) |  |
| <70% | 199 (38) | 144 (37) | 55 (41) |  |
| Unknown | 14 | 12 | 2 |  |
| **number of brain metastases, n (%)** |  |  |  | 0.062 |
| 1 | 149 (33) | 120 (36) | 29 (25) |  |
| 2 | 154 (34) | 115 (34) | 39 (34) |  |
| >2 | 150 (33) | 103 (30) | 47 (41) |  |
|  | Overall, N = 539<sup>1</sup> | <281 mg, N = 402 | ≥281 mg, N = 137 |  |
| Unknown | 86 | 64 | 22 |  |
| **extracran.met, n (%)** |  |  |  | 0.003 |
| no extracranial metastases | 288 (55) | 230 (59) | 58 (44) |  |
| extracranial metastases | 237 (45) | 162 (41) | 75 (56) |  |
| Unknown | 14 | 10 | 4 |  |
| **localization of brain metastases, n (%)** |  |  |  | 0.57 |
| supratentorial | 332 (63) | 244 (62) | 88 (65) |  |
| infratentorial | 90 (17) | 71 (18) | 19 (14) |  |
| supra- and infratentorial | 107 (20) | 79 (20) | 28 (21) |  |
| Unknown | 10 | 8 | 2 |  |
| **entity, n (%)** |  |  |  | 0.40 |
| breast cancer | 93 (17) | 71 (18) | 22 (16) |  |
| colorectal carcinoma | 8 (1.5) | 4 (1.0) | 4 (2.9) |  |
| melanoma | 91 (17) | 62 (15) | 29 (21) |  |
| NSCLC | 286 (53) | 218 (54) | 68 (50) |  |
| oesophageal carcinoma | 1 (0.2) | 1 (0.2) | 0 (0) |  |
| renal cell cancer | 26 (4.8) | 18 (4.5) | 8 (5.8) |  |
|  | Overall, N = 539<sup>1</sup> | <281 mg, N = 402 | ≥281 mg, N = 137 |  |
| SCLC | 33 (6.1) | 27 (6.7) | 6 (4.4) |  |
| unknown primary | 1 (0.2) | 1 (0.2) | 0 (0) |  |
| **Ki67, n (%)** |  |  |  | 0.93 |
| < 30 | 267 (58) | 199 (58) | 68 (58) |  |
| ≥ 30 | 195 (42) | 146 (42) | 49 (42) |  |
| Unknown | 77 | 57 | 20 |  |
| **PDL1i_TPS, n (%)** |  |  |  | 0.93 |
| < 30 | 102 (64) | 74 (64) | 28 (64) |  |
| ≥ 30 | 57 (36) | 41 (36) | 16 (36) |  |
| Unknown | 380 | 287 | 93 |  |
| **tumor.volume, n (%)** |  |  |  | 0.59 |
| < 10 | 146 (43) | 109 (43) | 37 (40) |  |
| ≥ 10 | 197 (57) | 142 (57) | 55 (60) |  |
| Unknown | 196 | 151 | 45 |  |
| **edema.volume, n (%)** |  |  |  | 0.091 |
| < 50 | 151 (46) | 118 (49) | 33 (38) |  |
| ≥ 50 | 176 (54) | 123 (51) | 53 (62) |  |
| Unknown | 212 | 161 | 51 |  |
|  | Overall, N = 539<sup>1</sup> | <281 mg, N = 402 | ≥281 mg, N = 137 |  |

<sup>1</sup> Median (IQR); n (%)
<sup>2</sup> Wilcoxon rank sum test; Pearson's Chi-squared test; Fisher's exact test

**Table 1b:** Comparison of the two patient groups of interest before propensity score matching within the whole patient cohort (“3months and KPS<50excl”) (N=483). The optimal cutpoint value of 281 mg for perioperative dexamethasone was used to dichotomize patients.

| Characteristic | Treatment |  |  | p-value<sup>2</sup> |
| --- | --- | --- | --- | --- |
|  | Overall, N = 483<sup>1</sup> | <294 mg, N = 375 | ≥294 mg, N = 108 |  |
| age (years), Median (IQR) | 64 (54 – 71) | 64 (54 – 71) | 65 (55 – 70) | >0.99 |
| gender, n (%) |  |  |  | 0.70 |
| female | 256 (53) | 197 (53) | 59 (55) |  |
| male | 227 (47) | 178 (47) | 49 (45) |  |
| pre-operative dexa (mg), Median (IQR) | 80 (24 – 144) | 60 (24 – 120) | 240 (168 – 336) | <0.001 |
| post-operative dexa (mg), Median (IQR) | 94 (62 – 132) | 84 (60 – 112) | 150 (102 – 230) | <0.001 |
| peri-operative dexa (mg), Median (IQR) | 187 (132 – 276) | 164 (114 – 207) | 387 (336 – 452) | <0.001 |
| KPS_group, n (%) |  |  |  | 0.49 |
| ≥70% | 306 (65) | 240 (66) | 66 (62) |  |
| <70% | 164 (35) | 124 (34) | 40 (38) |  |
| Unknown | 13 | 11 | 2 |  |
| number of brain metastases, n (%) |  |  |  | 0.093 |

| Characteristic | Treatment |  |  |  |
| --- | --- | --- | --- | --- |
|  | Overall, N = 483<sup>1</sup> | <294 mg, N = 375 | ≥294 mg, N = 108 | p-value<sup>2</sup> |
| 1 | 133 (33) | 110 (35) | 23 (26) |  |
| 2 | 138 (34) | 109 (35) | 29 (33) |  |
| >2 | 131 (33) | 94 (30) | 37 (42) |  |
| Unknown | 81 | 62 | 19 |  |
| **extracran.met, n (%)** |  |  |  | 0.008 |
| no extracranial metastases | 270 (57) | 222 (61) | 48 (46) |  |
| extracranial metastases | 200 (43) | 144 (39) | 56 (54) |  |
| Unknown | 13 | 9 | 4 |  |
| **localization of brain metastases, n (%)** |  |  |  | 0.25 |
| supratentorial | 298 (63) | 225 (61) | 73 (69) |  |
| infratentorial | 76 (16) | 64 (17) | 12 (11) |  |
| supra- and infratentorial | 100 (21) | 79 (21) | 21 (20) |  |
| Unknown | 9 | 7 | 2 |  |
| **entity, n (%)** |  |  |  | 0.54 |
| breast cancer | 88 (18) | 69 (18) | 19 (18) |  |
| colorectal carcinoma | 7 (1.4) | 4 (1.1) | 3 (2.8) |  |
| melanoma | 75 (16) | 53 (14) | 22 (20) |  |
| NSCLC | 261 (54) | 207 (55) | 54 (50) |  |
| oesophageal carcinoma | 1 (0.2) | 1 (0.3) | 0 (0) |  |
| renal cell cancer | 24 (5.0) | 18 (4.8) | 6 (5.6) |  |
| SCLC | 26 (5.4) | 22 (5.9) | 4 (3.7) |  |
| unknown primary | 1 (0.2) | 1 (0.3) | 0 (0) |  |
| **Ki67, n (%)** |  |  |  | 0.49 |
| < 30 | 239 (58) | 183 (57) | 56 (61) |  |
| ≥ 30 | 175 (42) | 139 (43) | 36 (39) |  |
|  | Overall, N = 483<sup>1</sup> | <294 mg, N = 375 | ≥294 mg, N = 108 | p-value<sup>2</sup> |
| Unknown | 69 | 53 | 16 |  |
| **PDL1i_TPS, n (%)** |  |  |  | 0.63 |
| < 30 | 96 (68) | 76 (69) | 20 (65) |  |
| ≥ 30 | 45 (32) | 34 (31) | 11 (35) |  |
| Unknown | 342 | 265 | 77 |  |
| **tumor.volume, n (%)** |  |  |  | 0.87 |
| < 10 | 130 (42) | 99 (41) | 31 (42) |  |
| ≥ 10 | 182 (58) | 140 (59) | 42 (58) |  |
| Unknown | 171 | 136 | 35 |  |
| **edema.volume, n (%)** |  |  |  | 0.13 |
| < 50 | 138 (46) | 112 (49) | 26 (38) |  |
| ≥ 50 | 160 (54) | 118 (51) | 42 (62) |  |
| Unknown | 185 | 145 | 40 |  |

<sup>1</sup> Median (IQR); n (%)
<sup>2</sup> Wilcoxon rank sum test; Pearson's Chi-squared test; Fisher's exact test

### 2.3 Cutpoints for perioperative Dexamethasone dosage regimens and association with overall survival

Next, cutpoint for cumulative perioperative dexamethasone (i.e. pre-operative, post-operative as well as pre- and postoperative or total) were evaluated in terms of its association as a predictor variable for OS. Cutpoint values for COHORT A and B in the pre-, postoperative phase are given in table 1a-c. As for a cutpoint of cumulative dexamethasone in the pre-operative period for COHORT A (40 mg) and COHORT B (184 mg) there were significant differences in median OS between patients with < 40 mg and ≥40 mg and < 184 mg and ≥184, respectively (**eFigure 3b in the online supplements**). In contrast, only the cutpoint for postoperative dexamethasone dosage for COHORT A (180 mg) showed a significant association with OS, whereas for COHORT B the established cutpoint (180 mg) did not result in a significant difference in OS when comparing two dichotomized patient groups (**eFigure 3b in the online supplements**). As for the cumulative dexamethasone dose in the pre- and post-operative period patients of COHORT A were dichotomized into two groups each: patients with < 281 mg showing a median OS of 12.9 months (95% CI: 11.0-15.2) vs. those patients with ≥281 mg cumulative dexamethasone in the pre- and postoperative period with an OS of 8.43 months (95% CI: 5.6-10.9) (p=0.0012). For COHORT B optimal cutpoint for the dexamethasone dose in the pre- and postoperative period was 294 mg of post-operative dexamethasone: patients with < 294 mg cumulative dexamethasone showed a median OS of 14.7 months (95% CI: 12.90-18.2) vs. patients with ≥294 mg cumulative dexamethasone in the post-operative period with an OS of 10.6 months (95% CI: 8.43-13.6) (p=0.0086) (**Figure 2, 3**). Additionally, we performed a 1:1 propensity score matching (PSM) for covariate balancing for the total cohort of patients (COHORT A), which resulted in COHORT C with 131 matched patients per group. Here, patients that received <281 mg cumulative dexamethasone in the perioperative period had a significantly higher OS (13.5 months; 95% CI: 10.9-17.9) than those patients that received ≥281 mg perioperative dexamethasone that showed an OS of 8.3 months (95% CI: 5.37-10.9) (p=0.0072) (**eFigure 4, 5 in the online supplements**).

**Table 1c:**
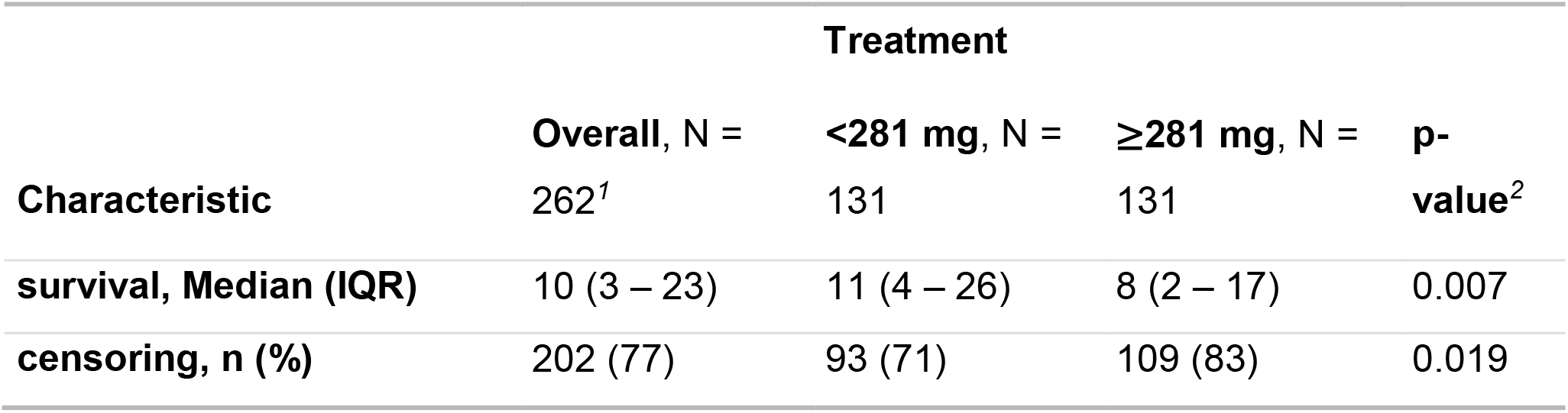
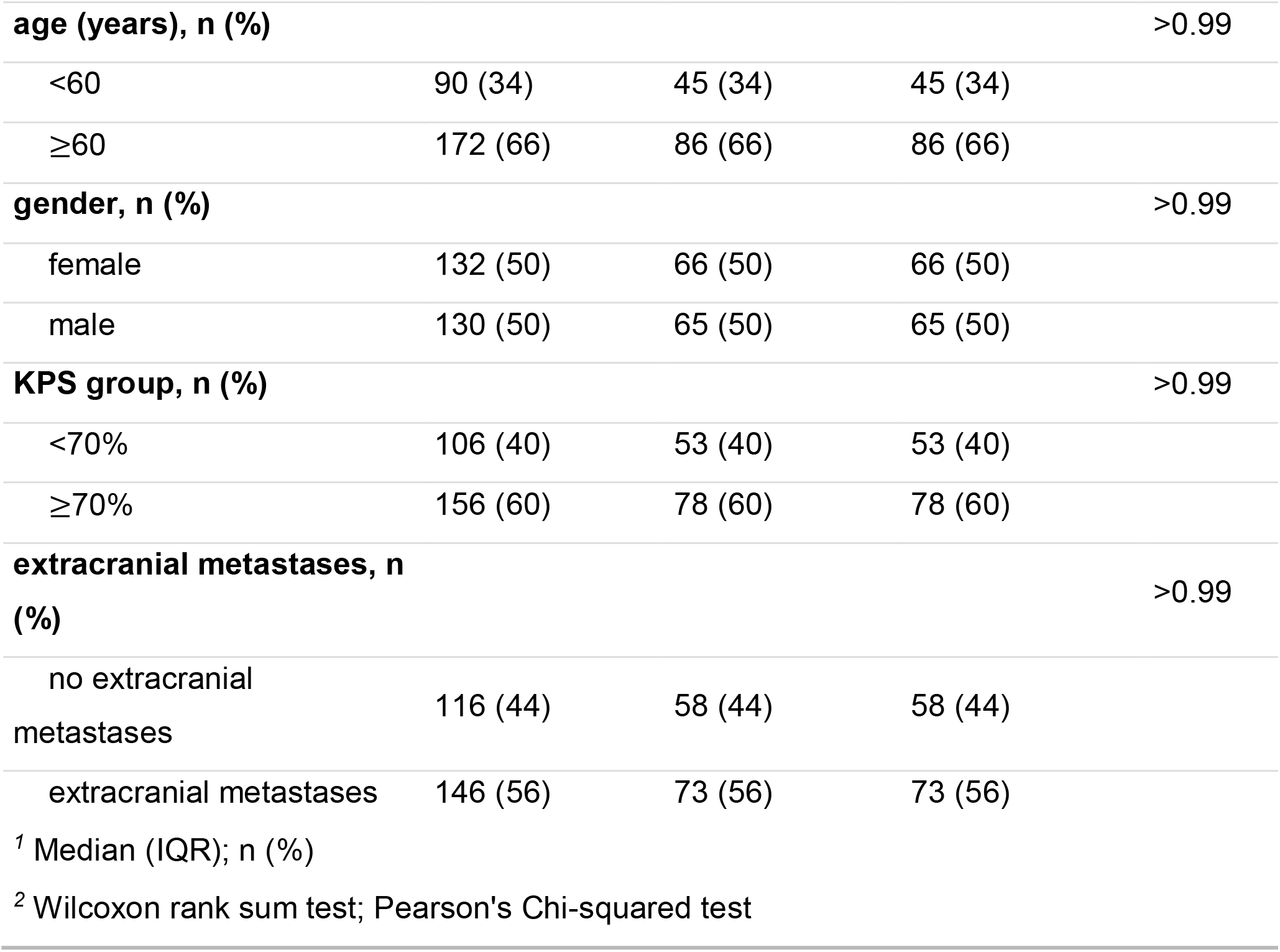
Summary of balance for matched data after performing propensity score matching for COHORT A produced COHORT C. Comparison of covariates after PSM for the whole patient cohort (n=539) with patients dichotomized according to the optimal of 281 mg (patients were grouped in those with <281 mg dexamethasone vs. ≥281 mg dexamethasone in the total perioperative phase (i.e. pre-operative and post-operative period). In total 216 number of observations were 1:1 matched; standard mean differences for unmatched data and matched data are plotted in **eFigure 4 in the supplements**.

**Figure 2:**
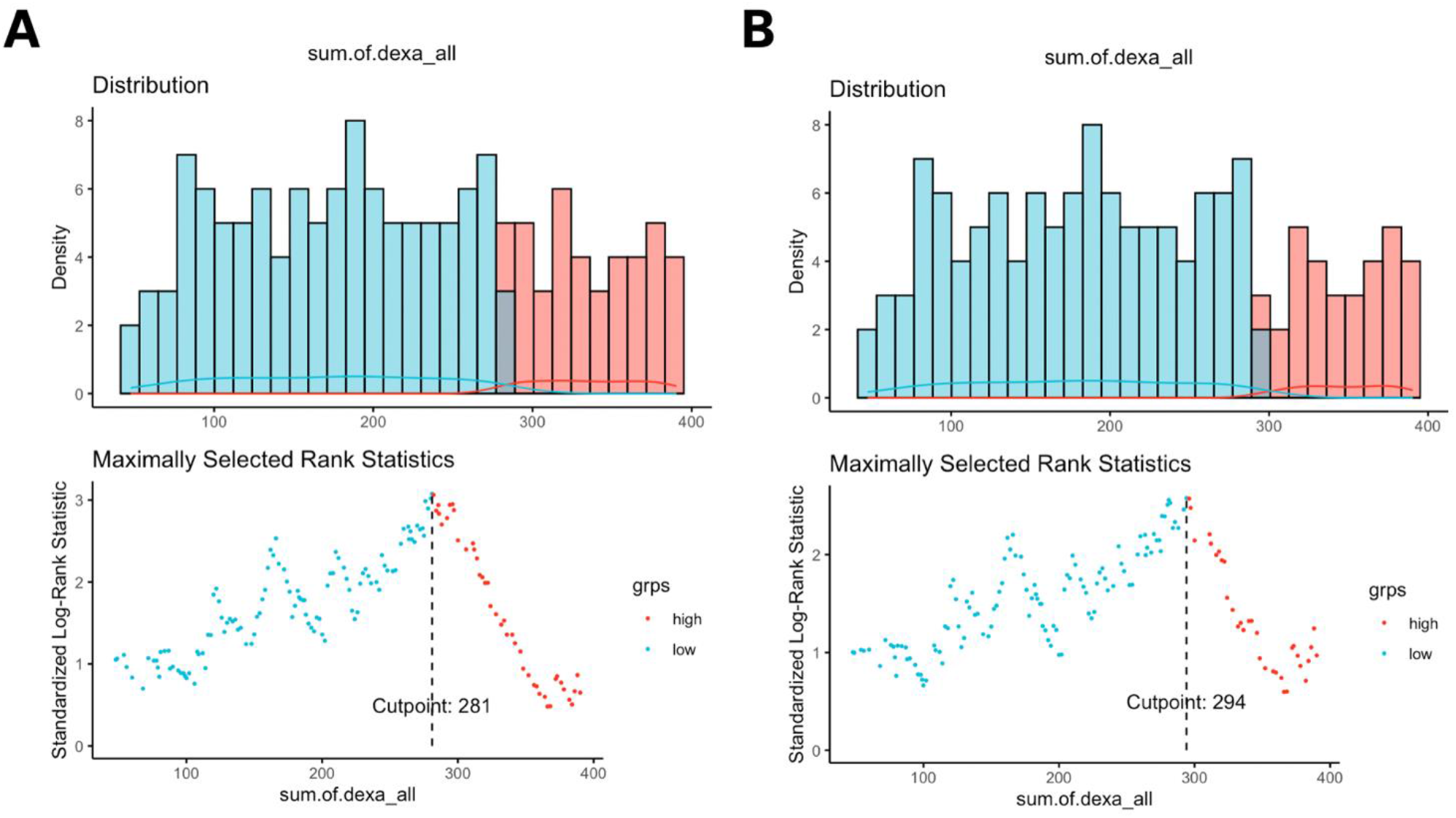
Cutpoint selection using maximally selected rank statistics in COHORT A displayed in **A**, with an optimal cutpoint for perioperative dexamethasone (pre-operative plus post-operative cumulative dexamethasone dosage) of 281 mg and for the subcohort COHORT B with an optimal cutpoint for total or perioperative dexamethasone dosage of 294 mg (**B**).

**Figure 3:**
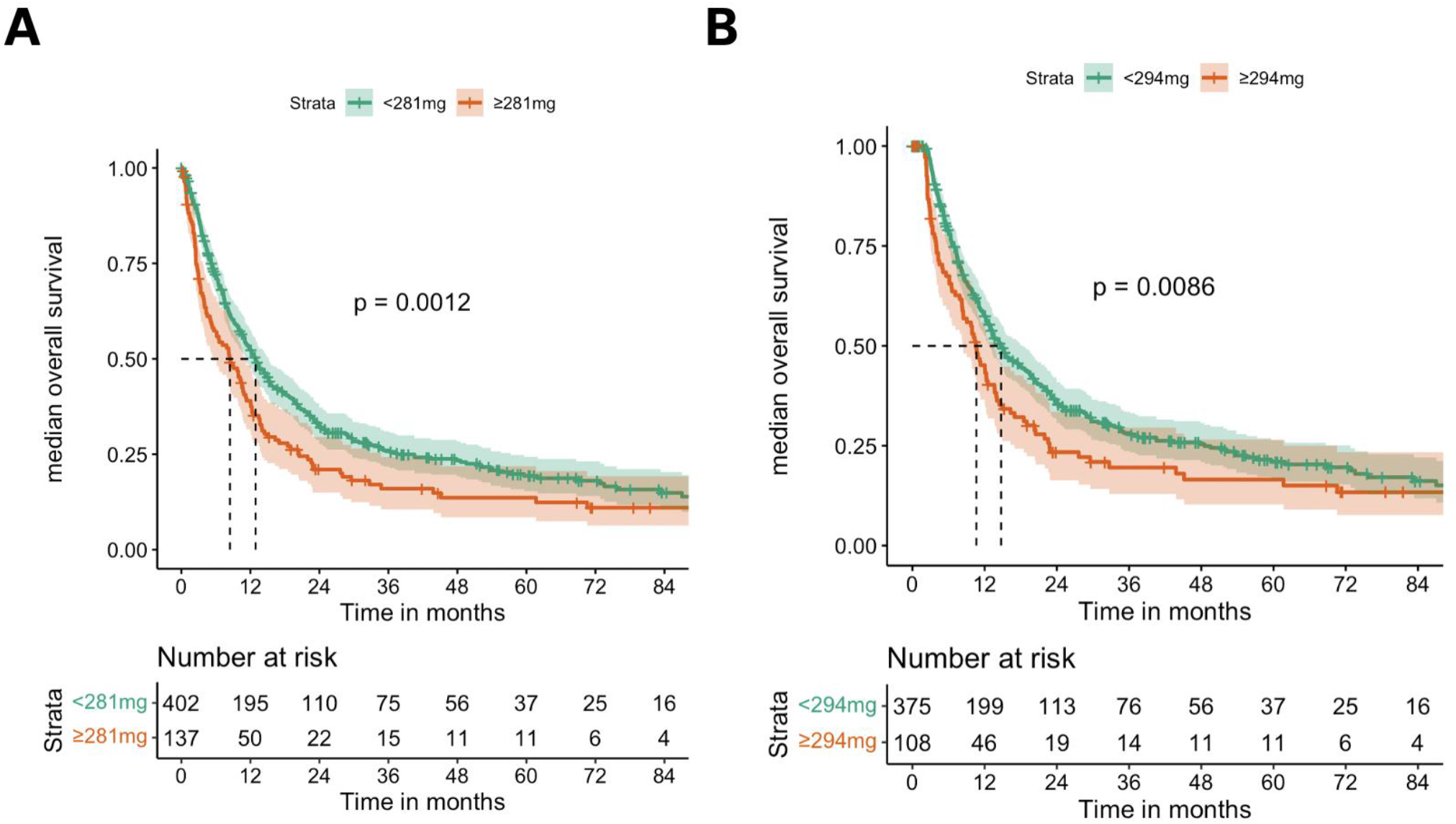
Kaplan-Meier curves and associated risk table displaying median overall survival of the 2 cohorts of interest COHORT A (“all”) (**A**) and the COHORT B (“3months and KPS<50excl”) (**B**).

### 2.4 Univariable and multivariable analysis to identify prognostic factors

On univariable analysis there was a statistically significant association between the cumulative perioperative dexamethasone dose and median OS in both cohorts COHORT A ([HR] 1.52, 95% CI: 1.25-1.85, p<0.001) and COHORT B ([HR] 1.38, 95% CI: 1.08-1.76, p=0.009). Multivariable Cox regression for perioperative dexamethasone and covariates which were considered as potential confounders (i.e. age, gender, KPS and presence of extracranial metastasis at baseline) showed that high perioperative (“total”) dexamethasone was interpedently associated with shorter OS in case of COHORT A ([HR] 1.47, 95% CI: 1.20-1.80, p<0.001) as well as the subgroup COHORT B ([HR] 1.35, 95% CI: 1.09-1.68, p=0.007) (**Table 2a**). Other independent prognostic factors for both cohorts were age and KPS (**Table 2b**). Importantly, also for the matched dataset of patients within the whole patient cohort (COHORT C) stratified into patients with < 281 mg perioperative dexamethasone and patients with ≥281 mg cumulative dexamethasone in the pre- and postoperative period showed that dexamethasone was an independent prognostic factor associated with OS ([HR] 1.46, 95% CI: 1.11-1.92, p=0.012) (**Table 2c**). Mean standard differences are displayed in **eFigure 3 in the supplements.**

**Table 2a:** Univariable and multivariable Cox proportional regression in the whole patient cohort. Cox regression analysis of potential prognostic covariates and overall survival for the whole patient cohort (“all”) (N=539).

|  |  |  |  |  |
| --- | --- | --- | --- | --- |
| Dependent:<br>Surv(survival,<br>censoring) |  | all | HR (CPH univar. analysis) | HR (CPH<br>multivar.analysis) |
| age, Mean (SD) |  | 63.0<br>(12.1) | 1.02 (1.01-1.03, p<0.001) | **1.02 (1.01-1.03, p<0.001)** |
| gender | female | 279<br>(51.8) | - | - |
|  | male | 260<br>(48.2) | 1.20 (0.99-1.46, p=0.061) | - |
| KPS, Mean (SD) |  | 78.6<br>(14.4) | 0.97-0.98, p<0.001 | **0.97-0.99, p<0.001** |
| extracran.met<br>(extracranial<br>metastasis) | Not<br>present | 288<br>(54.9) | - | - |
|  | Present | 237<br>(45.1) | 1.43 (1.15-1.77, p=0.001) | **1.32 (1.05-1.64, p=0.016)** |
| cut_off_before_and<br>_after_281 (total<br>perioperative<br>dexamethasone) | <281 mg | 402<br>(74.6) | - | - |
|  | ≥ 281 mg | 137<br>(25.4) | 1.52 (1.25-1.85, p<0.001) | **1.47 (1.20-1.80, p<0.001)** |

**Table 2b:**
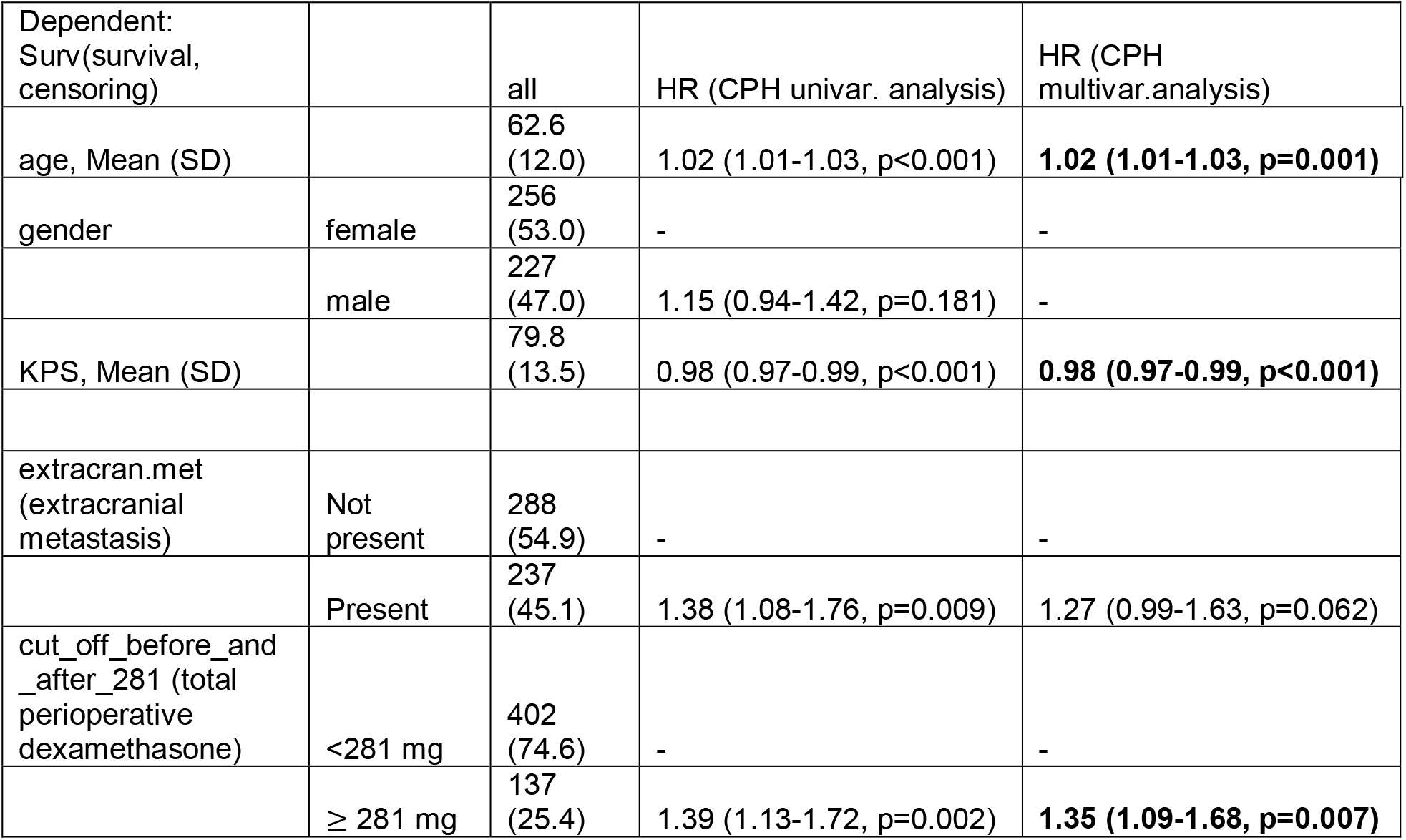
Univariable and multivariable Cox proportional regression in the subcohort “3months and KPS<50excl”. Cox regression analysis of potential prognostic covariates and overall survival for the patient cohort “3months and KPS<50excl” cohort (n=483).

| Dependent:<br>Surv(survival,<br>censoring) |  | all | HR (CPH univar. analysis) | HR (CPH<br>multivar.analysis) |
| --- | --- | --- | --- | --- |
| age, Mean (SD) |  | 62.6<br>(12.0) | 1.02 (1.01-1.03, p<0.001) | **1.02 (1.01-1.03, p=0.001)** |
| gender | female | 256<br>(53.0) | - | - |
|  | male | 227<br>(47.0) | 1.15 (0.94-1.42, p=0.181) | - |
| KPS, Mean (SD) |  | 79.8<br>(13.5) | 0.98 (0.97-0.99, p<0.001) | **0.98 (0.97-0.99, p<0.001)** |
| extracran.met<br>(extracranial<br>metastasis) | Not<br>present | 288<br>(54.9) | - | - |
|  | Present | 237<br>(45.1) | 1.38 (1.08-1.76, p=0.009) | 1.27 (0.99-1.63, p=0.062) |
| cut_off_before_and<br>_after_281 (total<br>perioperative<br>dexamethasone) | <281 mg | 402<br>(74.6) | - | - |
|  | ≥ 281 mg | 137<br>(25.4) | 1.39 (1.13-1.72, p=0.002) | **1.35 (1.09-1.68, p=0.007)** |

**Table 2c:** Univariable and multivariable Cox proportional regression in the matched patient cohort after propensity score matching. Univariable and multivariable Cox proportional regression analysis for matched data for the whole patient cohort (“all”) (N=539). See also **eFigure 3 and 4**.

| Dependent:<br>Surv(survival,<br>censoring) |  | all | HR (CPH univar. analysis) | HR (CPH<br>multivar.analysis) |
| --- | --- | --- | --- | --- |
| age, Mean (SD) | <60 | 90<br>(34.4) | - | - |
|  | ≥60 | (65.6) | 1.48 (1.10-2.00, p=0.009) | **1.58 (1.17-2.14, p=0.003)** |
| gender | female | 132<br>(50.4) | - | - |
|  | male | 130<br>(49.6) | 0.95 (0.72-1.25, p=0.706) | - |
| KPS, Mean (SD) | <70% | 106<br>(40.5) | - | - |
|  | ≥70% | 156<br>(59.5) | 0.67 (0.51-0.89, p=0.005) | **0.67 (0.51-0.89, p=0.005)** |
| extracran.met<br>(extracranial<br>metastasis) | Not<br>present | 116<br>(44.3) | - | - |
|  | Present | 146<br>(55.7) | 1.45 (1.09-1.91, p=0.010) | **1.54 (1.16-2.05, p=0.003)** |
| cut_off_before_and<br>_after_281 (total<br>perioperative<br>dexamethasone) | <281 mg | 402<br>(74.6) | - | - |
|  | ≥ 281 mg | 137<br>(25.4) | 1.46 (1.11-1.92, p=0.007) | **1.43 (1.08-1.88, p=0.012)** |

## DISCUSSION/CONCLUSION

Corticosteroids such as dexamethasone are a standard drug in the treatment of patients with brain metastases. They are used to control the mass effect of perifocal tumor edema and the resulting neurologic deficitsts^12^. Treatment recommendations in this context lack evidence and are primarily based on expert opinion and few retrospective data^3, 4, 10^. With advent of immunotherapy or CPI dexamethasone dosage and down-taper schemes will likely gain importance as for other entities or situations. However, one randomized study with patients with brain metastases receiving either 4 mg, 8 mg or 16 mg dexamethasone daily reported improvement of symptoms by means of KPS at day 7 and day 28, respectively^18^. Importantly, there is a marked variability between centers and treating physicians in terms of dexamethasone recommendations (e.g. initiation and down tapering)^9, 12, 19^. In this retrospective, comparative effectiveness study we characterize the dosage of dexamethasone and its impact on survival in patients that underwent brain metastasis resection. Here, a cutpoint estimation method was used to evaluate optimal value selection for survival prediction of pre-operative, postoperative and total dexamethasone. Pre-operative dexamethasone, but not postoperative dexamethasone showed an association to OS in the whole cohort of patients with resected brain metastases. Maximally selected rank statistics were performed for pre-operative, postoperative and total (pre- and postoperative) dexamethasone dosage with the total dexamethasone displaying the strongest association with OS resulting in a cut-point of 281 mg. Patients receiving a high total cumulative dexamethasone dose (i.e., patients receiving ≥281 mg) showed significantly reduced OS-an effect that was confirmed in a subgroup analysis of patients after applying exclusion criteria (i.e., exclusion of patients with short-term survival < 2 months or patients with postoperative KPS < 50%) **(Figure 2, 3**). These observations could be further confirmed by multivariable Cox regression analysis for unmatched data as well as for matched data after performing PSM where dexamethasone with a cut-point of 281 mg in the total cohort was significantly associated with an increased hazard for death (**Figure 2, 3**). In general, our observations are in line with previously published research in the context of perioperative dexamethasone in Glioblastoma patients. Here, the sum of cumulative dexamethasone doses in the pre-operative and post-operative period seems to have impact on OS as reported previously. For instance, Zhou et al. demonstrated in a single-center PSM study that postoperative day (POD) 0 to 21 dexamethasone dosage correlated with overall survival^17^. Interestingly, Medikonda et al. recently showed that that pre-operative, but not post-operative or pre- and postoperative Dexamethasone dosage in Glioblastoma patients was linked to shorter OS and reported a hazard ratio of 3.0 (95% CI: 0.9-9.4). In contrast, the effect of combined pre- and post-operative dexamethasone on survival was less pronounced in our cohort [HR] 1.47, 95% CI: 1.20-1.80, p<0.001). Additionally, upon stratification into subgroups of dexamethasone intake (cumulative pre-operative, postoperative and perioperative (total) dexamethasone intake), we only observed total dexamethasone to be associated with OS. Interestingly, there was no significant correlation between cumulative dexamethasone dose and tumor volume or edema volume (**eFigure 2a in the supplements**), which is in contrast to the observation from Zhou and colleagues for glioblastoma^17^. Limitations of this study include the retrospective nature of the data originating from a retrospective and prospective brain metastasis registry. In contrast to other studies, we mainly focused on inpatient records to assure data quality although insufficient data capture and entry in patient files can lead to bias as those patients with insufficient documentation of dexamethasone doses were excluded (**Figure 1**). Although we performed a PSM for prognostic clinical covariates such as age, and KPS as well as extracranial disease burden, there are certainly other confounding factors we do not consider into the matching process in this study, which include tumor volume and edema volume. These however are of prognostic value and were not considered into the matching process. Additionally, larger patient cohorts from multicenter registries with standardized documentation of dexamethasone dosage would be necessary to further investigate the role of dexamethasone and its impact on patient outcome. This study consists only of surgical patients which tend to be clinically symptomatic. Therefore, patients that did not undergo surgery were excluded from this study. Our study has potential implications with respect to the efficacy of CPI in patients with brain metastases that may be candidates to receive CPIs after brain metastasis resection and RTx^20-23^. In this regard in patients with metastatic disease several retrospective studies indicated that timing of initiation of corticosteroids may influence therapy response to CPI; for example, Maslov and colleagues showed that patients treated with corticosteroids within 2 months of initiation with CPI showed significantly shorter OS than patients receiving corticosteroids after 2 months of initiation of treatment with CPIs^22^. Future prospective, randomized-controlled and interdisciplinary studies should systematically evaluate the benefit and complications or impact on OS but also on progression-free survival according to the Response Assessment in Neuro-Oncology Brain Metastasis (RANO-BM) criteria in different tumor entities. Additionally, exploratory endpoints, serum and tissue-based biomarker studies of resected brain metastases pre-exposed to dexamethasone should be initiated and could aid in gaining insights into the changes of the systemic and local tumor immune microenvironment in these patients^24-26^. This would also be informative for future clinical trials recruiting patients with brain metastases since patients with brain metastases are finally becoming more and more included in clinical trials independent of the tumor type. These studies could help in better understanding the role of perioperative dexamethasone or corticosteroids in terms of patient outcome, complications, or predicting the impact on post-operative “adjuvant” treatment including upcoming immunotherapy or CPIs.

## Figures and Tables

See below to see the main figures and tables and separate word file including supplementary figures „2. supplementary figures and material_08.01.2022_1334PM_DW”.

## Data Availability

Raw data and R code and raw data will be made available at github upon request.

## Author Contributions

D. W., N. F., P.V. and J. O. designed and prepared the manuscript. D. W., J. B., R. P., Z. S., P. P., AG. K., C. J., A. T.-F., M. R., H. R., D. C., M. S., P. T., P. B., D. H., M. K., R. X., F. E., D. K., M. M., L. B., N. F., P. V. and J. O. discussed and reviewed the manuscript. D. W., J. B., R. P., Z. S., P. P., AG. K., C. J., A. F., AG. K., A. T.-F., N. F. and J. O. have performed collection and organization of data. D. W. and A. T.-F. analyzed the data. R. K. provided patient records from the archive of the Charité. H. R., D. C., P. B. and D. H., provided information on histopathology. All authors read and approved the final version of the manuscript.

## Acknowledgments

We thank Dr. med. Susen Burock for data queries and patient data from the Berlin tumor registry (Berliner Krebsregister). D.W., N. F. and J.O. had full access to all the data in the study and take responsibility for the integrity of the data and accuracy of the data analysis.

## Supplementary Material

See separate online content file including supplementary figures (eFigure 1-4) and tables (eTable 1a-d, eTable 2a-d).

## Data availability statement

The datasets and R code and the usage of the R packages supporting the conclusions of this study will be made available on Github by the authors and can be downloaded upon request.

