## supplementary material for "Impact of Perioperative Dexamethasone in the Context of Neurosurgical Brain Metastasis Resection"

**eTable 1a-d:** Demographic, clinical and radiologic, histopathological and treatment-related characteristics at baseline of COHORT A

**eTable 2a-d:** Demographic, clinical and radiologic, histopathological and treatment-related characteristics at baseline of COHORT B

**eFigure 1:** Kaplan Meier survival estimates and comparison of COHORT A with COHORT B

**eFigure 2a-b:** Correlation of total dexamethasone with clinical parameters and daily perioperative dexamethasone doses

**eFigure 3a-b:** Kaplan-Meier analysis for overall survival of different dexamethasone dosage patient groups in COHORT A and B

**eFigure 3:** Propensity score matching and resulting mean standardized differences

**eFigure 4:** Kaplan-Meier survival curve for matched patients (COHORT C) after propensity score matching

This supplementary material has been provided by the authors to give readers additional information about their work.

**Suppl. table 1a: General patient characteristics**

| Characteristic | N = 539 |
|---|---|
| **Age, Median (IQR)** | 64 (55 – 72) |
| **Gender, n (%)** | |
| female | 279 (52) |
| male | 260 (48) |
| **signs and symptoms at baseline (OP1), n (%)** | |
| headache | 67 (12) |
| sensory-motor symptoms | 113 (21) |
| seizures | 52 (9.6) |
| alterations in behaviour | 47 (8.7) |
| aphasia | 41 (7.6) |
| vertigo/dyscoordination | 94 (17) |
| headache, nausea, vomitus | 32 (5.9) |
| visual impairment | 32 (5.9) |
| during staining | 26 (4.8) |
| incidental or others | 35 (6.5) |
| **time of brain metastasis resection, n (%)** | |
| brain metastasis resection after 1st of Januar 2015 | 329 (61) |
| brain metastasis resection before 1st of Januar 2015 | 210 (39) |
| **KPS after operation, n (%)** | |
| high | 326 (62) |
| low | 199 (38) |
| Unknown | 14 |
| **underlying other diseases, n (%)** | |
| no underyling other diseases | 124 (55) |
| underyling other diseases | 102 (45) |
| Unknown | 313 |

**eTable 1a: Demographic, clinical and radiologic characteristics at baseline**

General patient characteristics at baseline (i.e at time of first brain metastasis resection) of the total cohort of patients of interest (COHORT A) in this study for which further downstream analysis was performed (N=539). A high postoperative Karnofsky performance index (KPS) was defined as 70% or higher, whereas a KPS for lower than 70% was considered low. In 14 patients postoperative KPS and in 313 cases status regarding other diseases was not available or documented. The *gtsummary* package and windows word were used to describe tabular data of our patient cohort, including continuous, categorical or numerical variables.

**Suppl. table 1b: Radiological patient characteristics**

| **Characteristic** | **N = 539** |
|---|---|
| **number of brain metastases at baseline, n (%)** | |
| one | 149 (33) |
| two | 154 (34) |
| three or more | 150 (33) |
| Unknown | 86 |
| **anatomical site at at baseline, n (%)** | |
| frontal | 178 (33) |
| temporal | 92 (17) |
| parietal | 67 (13) |
| occipital | 59 (11) |
| cerebellar | 133 (25) |
| other | 6 (1.1) |
| Unknown | 4 |
| **presence of extracranial metastasis, n (%)** | |
| no presence of extracranial metastases at baseline | 288 (55) |
| presence of extracranial metastases at baseline | 237 (45) |
| Unknown | 14 |

**Suppl. table 1b: Radiological patient characteristics**

| Characteristic | N = 539 |
|---|---|
| **hemorrhage at baseline, n (%)** | |
| no hemorrhage at baseline | 389 (76) |
| hemorrhage at baseline | 124 (24) |
| Unknown | 26 |
| **hydrocephalus at baseline, n (%)** | |
| no hydrocephalus at baseline | 434 (87) |
| hydrocephalus at baseline | 63 (13) |
| Unknown | 42 |
| **localization of dominant brain metastasis, n (%)** | |
| supratentorial | 332 (63) |
| infratentorial | 90 (17) |
| both supra-/infratentorial | 107 (20) |
| Unknown | 10 |
| **tumor volume, n (%)** | |
| < 10 | 146 (43) |
| >/= 10 | 197 (57) |
| Unknown | 196 |
| **edema volume, n (%)** | |
| < 50 | 151 (46) |
| >/= 50 | 176 (54) |
| Unknown | 212 |

**eTable 1b:** Baseline radiological characteristics of the total COHORT A (N=539). For patients where no pre-operative MRI was available no exact annotation of anatomical site and no exact number of brain metastases were able to be retrieved. Similarly in 14 cases no CT staging imaging was available in the setting of the brain metastasis resection. In 196 and 212 cases there was no information on tumor volume and edema volume, respectively.

47

**Suppl. table 1c: Histopathological and biomarker-related patient characteristics**

| Characteristic | N = 539 |
|---|---|
| **entity, n (%)** | |
| breast cancer | 93 (17) |
| colorectal carcinoma | 8 (1.5) |
| melanoma | 91 (17) |
| NSCLC | 286 (53) |
| oesophageal carcinoma | 1 (0.2) |
| renal cell cancer | 26 (4.8) |
| SCLC | 33 (6.1) |
| unknown primary | 1 (0.2) |
| **histological subtype, n (%)** | |
| adenocarcinoma | 1 (0.2) |
| adenocarcinoma (breast) | 92 (17) |
| adenocarcinoma (colorectal) | 8 (1.5) |
| adenocarcinoma (lung) | 247 (46) |
| adenosquamous | 3 (0.6) |
| clear cell renal cell carcinoma | 26 (4.8) |
| melanoma | 91 (17) |
| metaplastic carcinoma | 1 (0.2) |
| neuroendocrine-differentiated tumor | 42 (7.8) |
| pulmonary sarcomatoid carcinoma | 1 (0.2) |
| squamous cell carcinoma (lung) | 26 (4.8) |
| undifferentiated | 1 (0.2) |
| **EGFR mutational status, n (%)** | 14 (7.3) |
| Unknown | 347 |
| **ALK status, n (%)** | 6 (3.4) |
| Unknown | 361 |

**Suppl. table 1c: Histopathological and biomarker-related patient characteristics**

| Characteristic | N = 539 |
|---|---|
| **ROS1 status, n (%)** | 0 (0) |
| Unknown | 375 |
| **TTF1 status, n (%)** | |
| negative | 104 (33) |
| positive | 214 (67) |
| Unknown | 221 |
| **progesterone receptor expression (Remmele score), n (%)** | |
| < 4 | 85 (97) |
| >/= 4 | 3 (3.4) |
| Unknown | 451 |
| **estrogen receptor expression (Remmele score), n (%)** | |
| < 4 | 60 (68) |
| >/= 4 | 28 (32) |
| Unknown | 451 |
| **Her2/NEU expression (DAKO score), n (%)** | |
| 0+ | 22 (25) |
| 1+ | 14 (16) |
| 2+ | 9 (10) |
| 3+ | 42 (48) |
| Unknown | 452 |
| **Ki67 index (%), n (%)** | |
| < 30 | 267 (58) |
| >/= 30 | 195 (42) |
| Unknown | 77 |
| **intracranial PD-L1 TPS (%), n (%)** | |
| < 1 | 102 (64) |

**Suppl. table 1c: Histopathological and biomarker-related patient characteristics**

| Characteristic | N = 539 |
|---|---|
| >/= 1 | 57 (36) |
| Unknown | 380 |

**eTable 1c:** Histopathological characteristics of COHORT A (N=539). Histology is listed separately from entity to show distinct histopathological subtypes. Oncogenic drivers such as EGFR, ALK, ROS1 and BRAF were available only in corresponding entity, e.g. BRAF for melanoma or EGFR for lung cancer; the NA values were classified as not available or not applicable where needed to simplify the structure of the table. Similarly immune scores (Remmele score) for progesterone receptor (PR) and estrogen receptor (ER) as well as Her2/NEU2 score (DAKO score) were only evaluated and available in tissues from breast cancer patients. Ki67 index was dichotomized with a cut-off of 30%, whereas intracranial PD-L1 status (tumor proportion score, TPS) was dichotomized into samples with < 1% PD-L1 surface expression on tumor cell membranes, and those with ≥ 1% PD-L1 expression. Information on molecular characteristics (i.e. oncogenic driver mutations) or PD-L1 TPS of primary tumor tissue was not included into tabular data - only results from brain metastasis tissue are listed.

**Suppl. table 1d: Treatment-related patient characteristics**

| Characteristic | N = 539 |
|---|---|
| **primary tumor resection, n (%)** | |
| no primary tumor resection | 234 (45) |
| primary tumor resection | 281 (55) |
| Unknown | 24 |
| **naive, n (%)** | |
| naive | 289 (55) |
| pre-treated | 238 (45) |
| Unknown | 12 |

**Suppl. table 1d: Treatment-related patient characteristics**

| Characteristic | N = 539 |
|---|---|
| **adjuvant RTx, n (%)** | |
| no SRS | 206 (74) |
| SRS | 72 (26) |
| Unknown | 261 |
| **adjuvant systemic treatment, n (%)** | |
| BSC | 34 (8.8) |
| RTx only | 72 (19) |
| RTx + CTx | 142 (37) |
| RTx + targeted therapy | 38 (9.8) |
| RTx + CPI | 100 (26) |
| Unknown | 153 |

**eTable 1d:** Treatment-related characteristics of COHORT A. In 153 cases subsequent therapy after brain metastasis resection was not documented or incompletely documented, whereas 142 patients received radiation therapy (RTx) and chemotherapy (CTx) mainly including carboplatin- or cisplatin-based regimens, 100 patients checkpoint inhibition (CPI) mainly receiving combined chemoimmunotherapy (N=14), Nivolumab or Nivolumab/Ipilimumab combinations (N=29), or Pembrolizumab (N=35) or other combinations of different checkpoint inhibitors (N=22) (data not shown). 34 patients (8,8%) received best supportive care (BSC).

**Table 2a: General patient characteristics**

| Characteristic | N = 483 |
|---|---|
| **Age, Median (IQR)** | 64 (54 – 71) |
| **Gender, n (%)** | |
| female | 256 (53) |

**Table 2a: General patient characteristics**

| Characteristic | N = 483 |
| --- | --- |
| male | 227 (47) |
| **signs and symptoms at baseline (OP1), n (%)** | |
| headache | 65 (13) |
| sensory-motor symptoms | 97 (20) |
| seizures | 43 (8.9) |
| alterations in behaviour | 41 (8.5) |
| aphasia | 39 (8.1) |
| vertigo/dyscoordination | 84 (17) |
| headache, nausea, vomitus | 31 (6.4) |
| visual impairment | 31 (6.4) |
| during staining | 25 (5.2) |
| incidental or others | 27 (5.6) |
| **time of brain metastasis resection, n (%)** | |
| brain metastasis resection after 1st of Januar 2015 | 298 (62) |
| brain metastasis resection before 1st of Januar 2015 | 185 (38) |
| **KPS after operation, n (%)** | |
| high | 306 (65) |
| low | 164 (35) |
| Unknown | 13 |
| **underlying other diseases, n (%)** | |
| no underyling other diseases | 106 (52) |
| underyling other diseases | 96 (48) |
| Unknown | 281 |

**eTable 2a: Demographic, clinical and radiologic characteristics at baseline**

General patient characteristics at baseline (i.e at time of first brain metastasis resection) of COHORT B for which further downstream analysis was performed (N=483), see consorts diagram **Figure 1**. A high postoperative Karnofsky performance index (KPS) was defined as 70% or higher, whereas a KPS for lower than 70% was considered low. In 13 patients postoperative KPS was not available. The *gtsummary* package and windows word were used to describe tabular data of our patient cohort, including continuous, categorical or numerical variables.

**Table 2b: Radiological patient characteristics**

| **Characteristic** | **N = 483** |
|---|---|
| **number of brain metastases at baseline, n (%)** | |
| one | 133 (33) |
| two | 138 (34) |
| three or more | 131 (33) |
| Unknown | 81 |
| **anatomical site at at baseline, n (%)** | |
| frontal | 153 (32) |
| temporal | 84 (18) |
| parietal | 63 (13) |
| occipital | 55 (11) |
| cerebellar | 120 (25) |
| other | 4 (0.8) |
| Unknown | 4 |
| **presence of extracranial metastasis, n (%)** | |
| no presence of extracranial metastases at baseline | 270 (57) |
| presence of extracranial metastases at baseline | 200 (43) |
| Unknown | 13 |
| **hemorrhage at baseline, n (%)** | |

**Table 2b: Radiological patient characteristics**

| **Characteristic** | **N = 483** |
|---|---|
| no hemorrhage at baseline | 354 (77) |
| hemorrhage at baseline | 104 (23) |
| Unknown | 25 |
| **hydrocephalus at baseline, n (%)** | |
| no hydrocephalus at baseline | 388 (87) |
| hydrocephalus at baseline | 59 (13) |
| Unknown | 36 |
| **localization of dominant brain metastasis, n (%)** | |
| supratentorial | 298 (63) |
| infratentorial | 76 (16) |
| both supra-/infratentorial | 100 (21) |
| Unknown | 9 |
| **tumor volume, n (%)** | |
| < 10 | 130 (42) |
| >/= 10 | 182 (58) |
| Unknown | 171 |
| **edema volume, n (%)** | |
| < 50 | 138 (46) |
| >/= 50 | 160 (54) |
| Unknown | 185 |

**eTable 2b:** Baseline radiological characteristics of patients after excluding short-term survivors (< 2 months) and patients with very low KPS values (i.e. < 50%) (COHORT B) (N=483). For patients where no pre-operative MRI was available no exact annotation of anatomical site and no exact number of brain metastases were able to be retrieved. Similarly in 13 cases no CT staging imaging was available in the setting of the brain metastasis resection (**see also eTable1b in the supplementary section**).

**Table 2c: Histopathological and biomarker-related patient characteristics**

| Characteristic | N = 483 |
|---|---|
| **entity, n (%)** | |
| breast cancer | 88 (18) |
| colorectal carcinoma | 7 (1.4) |
| melanoma | 75 (16) |
| NSCLC | 261 (54) |
| oesophageal carcinoma | 1 (0.2) |
| renal cell cancer | 24 (5.0) |
| SCLC | 26 (5.4) |
| unknown primary | 1 (0.2) |
| **histological subtype, n (%)** | |
| adenocarcinoma | 1 (0.2) |
| adenocarcinoma (breast) | 87 (18) |
| adenocarcinoma (colorectal) | 7 (1.4) |
| adenocarcinoma (lung) | 227 (47) |
| adenosquamous | 3 (0.6) |
| clear cell renal cell carcinoma | 24 (5.0) |
| melanoma | 75 (16) |
| metaplastic carcinoma | 1 (0.2) |
| neuroendocrine-differentiated tumor | 36 (7.5) |
| pulmonary sarcomatoid carcinoma | 1 (0.2) |
| squamous cell carcinoma (lung) | 21 (4.3) |
| **EGFR mutational status, n (%)** | 14 (7.8) |
| Unknown | 304 |
| **ALK status, n (%)** | 6 (3.6) |

**Table 2c: Histopathological and biomarker-related patient characteristics**

| Characteristic | N = 483 |
|---|---|
| Unknown | 316 |
| **ROS1 status, n (%)** | 0 (0) |
| Unknown | 330 |
| **TTF1 status, n (%)** | |
| negative | 90 (31) |
| positive | 196 (69) |
| Unknown | 197 |
| **progesterone receptor expression (Remmele score), n (%)** | |
| < 4 | 80 (96) |
| >/= 4 | 3 (3.6) |
| Unknown | 400 |
| **estrogen receptor expression (Remmele score), n (%)** | |
| < 4 | 55 (66) |
| >/= 4 | 28 (34) |
| Unknown | 400 |
| **Her2/NEU expression (DAKO score), n (%)** | |
| 0+ | 20 (24) |
| 1+ | 14 (17) |
| 2+ | 9 (11) |
| 3+ | 40 (48) |
| Unknown | 400 |
| **Ki67 index (%), n (%)** | |
| < 30 | 239 (58) |
| >/= 30 | 175 (42) |
| Unknown | 69 |
| **intracranial PD-L1 TPS (%), n (%)** | |

**Table 2c: Histopathological and biomarker-related patient characteristics**

| Characteristic | N = 483 |
|---|---|
| < 1 | 96 (68) |
| >/= 1 | 45 (32) |
| Unknown | 342 |

**eTable 2c:** Histopathological characteristics of COHORT B (N=483). Histology is listed separately from entity to show distinct histopathological subtypes. Oncogenic drivers such as EGFR, ALK, ROS1 and BRAF were available only in corresponding entity, e.g. BRAF for melanoma or EGFR for lung cancer; the NA values were classified as not available or not applicable where needed to simplify the structure of the table. Similarly immune scores (Remmele score) for progesterone receptor (PR) and estrogen receptor (ER) as well as Her2/NEU2 score (DAKO score) were only evaluated and available in tissues from breast cancer patients. Ki67 index was dichotomized with a cut-off of 30% with 69 patients without documented Ki67 index. Information on molecular characteristics (i.e. driver mutations) of primary tumor tissue was not included into tabular data - only results from brain metastasis tissue were listed.

**Table 2d: Treatment-related patient characteristics**

| Characteristic | N = 483 |
|---|---|
| **primary tumor resection, n (%)** | |
| no primary tumor resection | 208 (45) |
| primary tumor resection | 253 (55) |
| Unknown | 22 |
| **naive, n (%)** | |
| naive | 260 (55) |
| pre-treated | 213 (45) |
| Unknown | 10 |

**Table 2d: Treatment-related patient characteristics**

| Characteristic | N = 483 |
|---|---|
| **adjuvant RTx, n (%)** | |
| no SRS | 192 (73) |
| SRS | 72 (27) |
| Unknown | 219 |
| **adjuvant systemic treatment, n (%)** | |
| BSC | 15 (4.4) |
| RTx only | 63 (18) |
| RTx + CTx | 135 (39) |
| RTx + targeted therapy | 38 (11) |
| RTx + CPI | 93 (27) |
| Unknown | 139 |

**eTable 2d:** Treatment-related characteristics of COHORT B; in 139 cases subsequent therapy after brain metastasis resection was not documented or incompletely documented, whereas 135 patients received radiation therapy (RTx) and chemotherapy (CTx) mainly including carboplatin- or cisplatin-based regimens, 93 patients checkpoint inhibition (CPI) mainly receiving combined chemoimmunotherapy (N=14), Nivolumab or Nivolumab/Ipilimumab combinations (N=29), or Pembrolizumab (N=35) or other combinations of different checkpoint inhibitors (N=22) (data not shown) (see also supplementary table 1d).

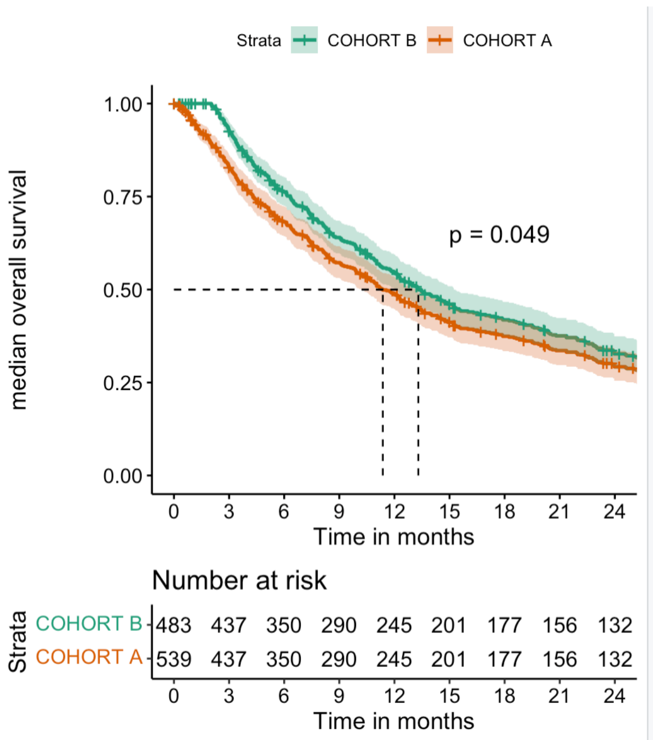


**eFigure 1: Kaplan Meier survival estimates and comparison of COHORT A with COHORT B**

Kaplan Meier survival analysis for OS in the entire patient cohort of brain metastasis patients subjected to craniotomy and brain metastasis removal (N=539) (“all” or COHORT A) with an OS of 11.4 months (95%CI: 10.2 – 13.2) in comparison to the collective of patients after applying exclusion criteria (i.e. excluding patients with less than 2 months OS and patients with KPS less than 50%, “2 months and KPS<50excl” or COHORT B) showing a OS of 13.3 months (95%CI: 12.1 – 15.2) (p=0.049).

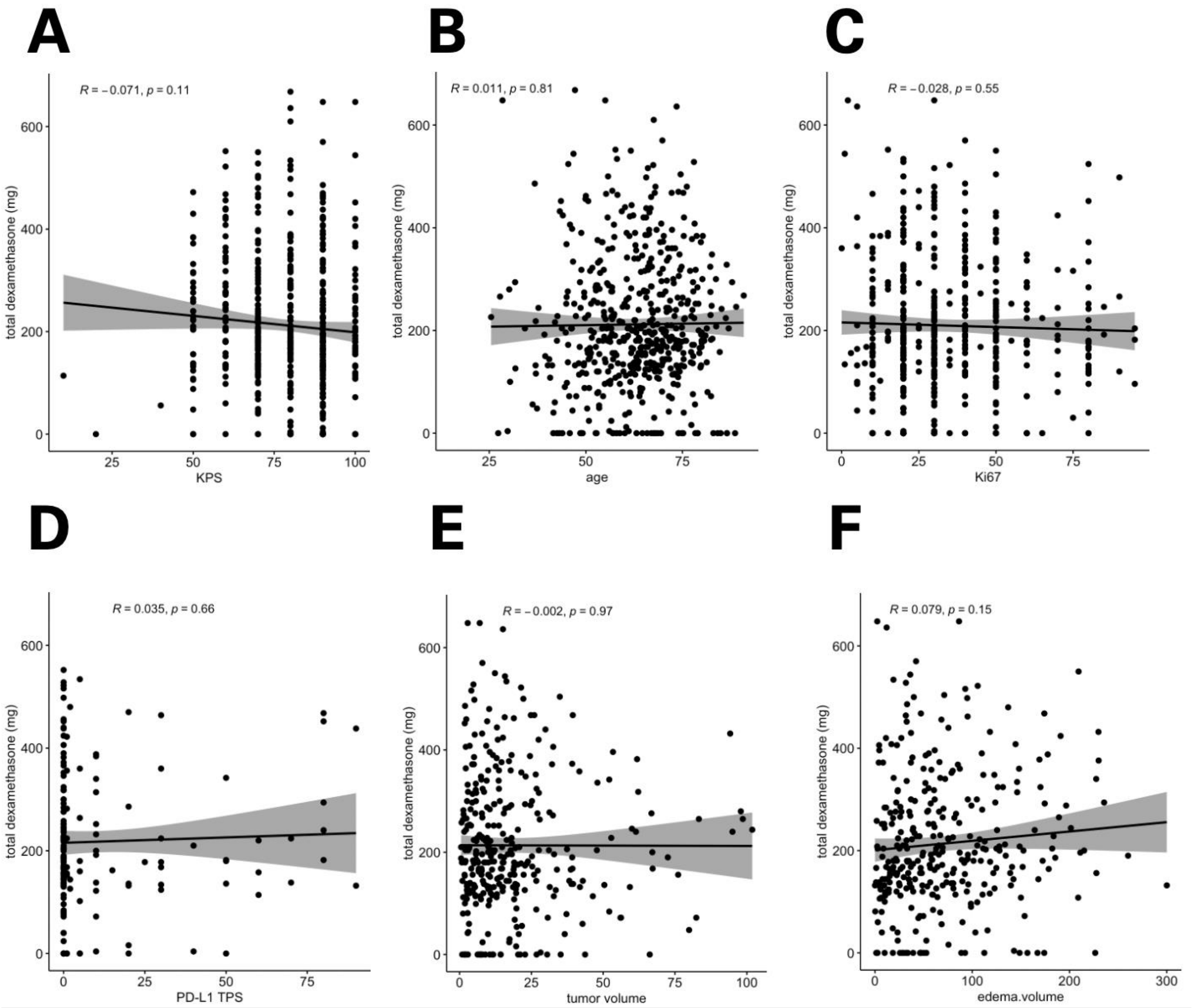


**eFigure 2a: Correlation of total dexamethasone with clinical parameters**

Pearson correlation (95% CI in gray; Pearson correlation coefficient R and cognate p value in the upper left section of each graphic) of total dexamethasone (i.e. pre-operative and post-operative dexamethasone) in resected brain metastases with a) age, b) KPS, c) Ki67 and d) PD-L1 TPS, e) tumor volume and f) edema volume. Only data for COHORT A are shown.

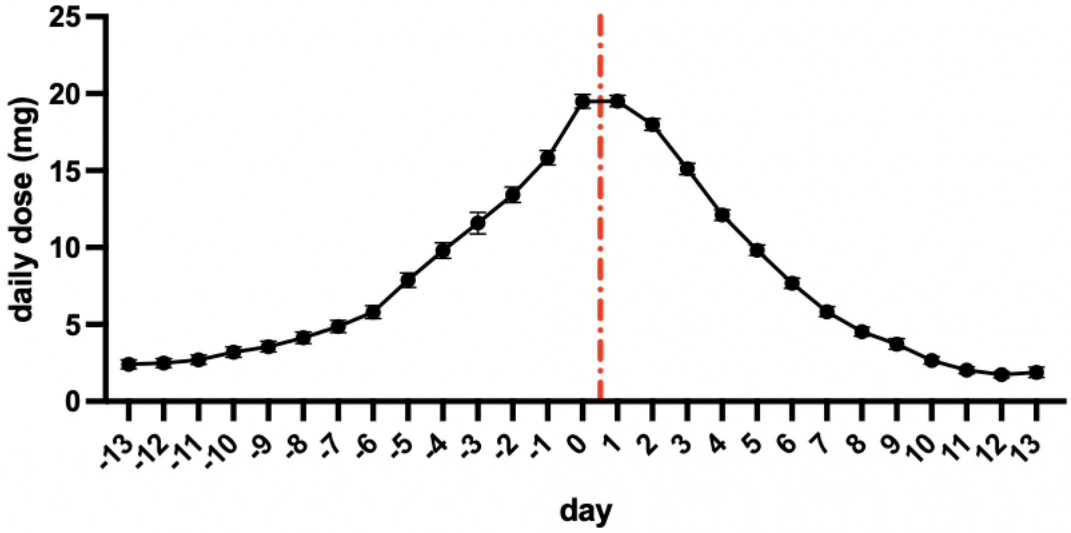


**eFigure 2b: Daily perioperative dexamethasone doses**

Mean values (mean with standard error of the means (SEM)) of daily dexamethasone for the total cohort of patients on the y-axis and days in relation to day 0 (day of surgical brain metastasis resection) are displayed for the total cohort of patients (N=539). Graph was produced with GraphPad Prism. Data for cohort A are shown.

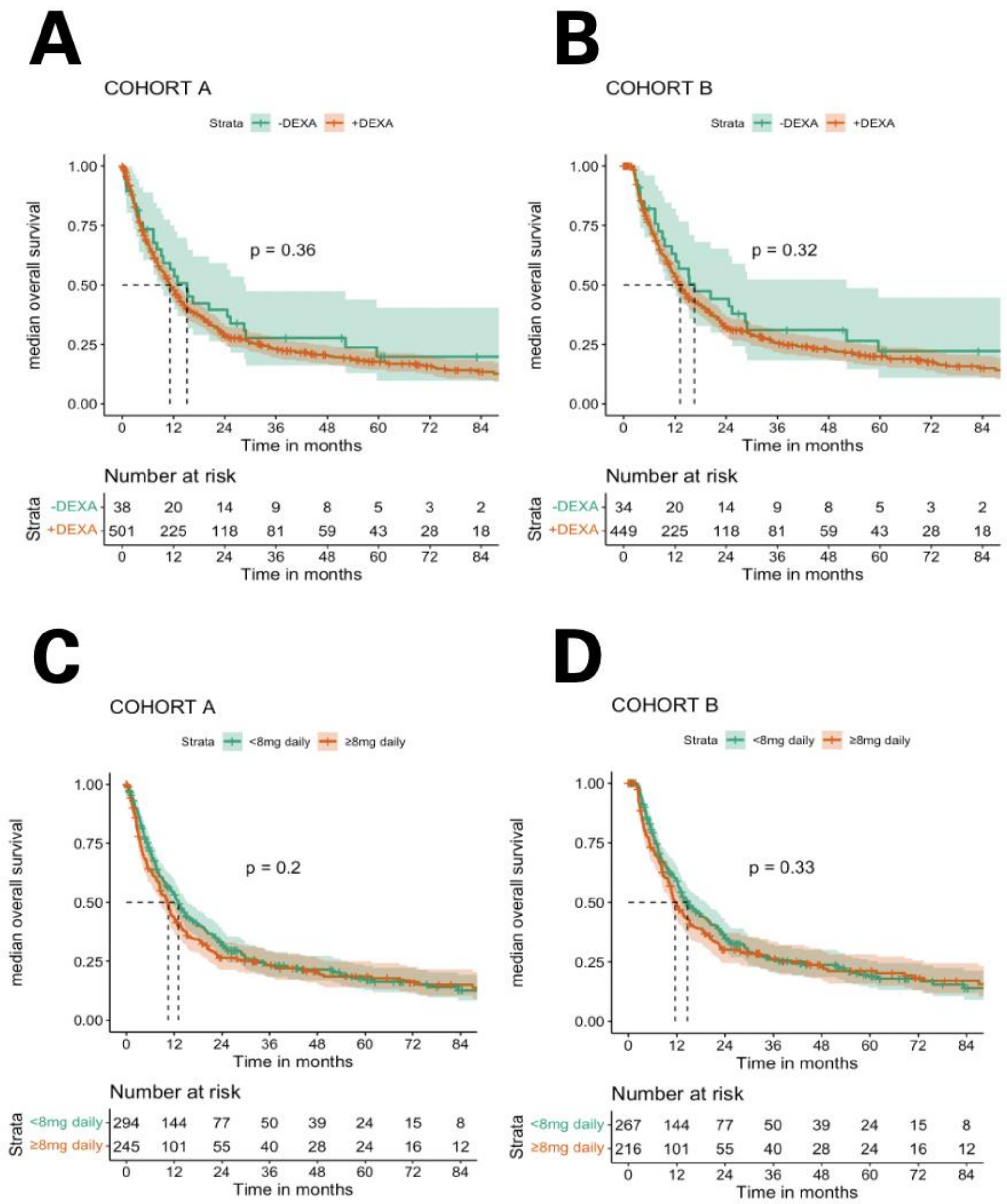


**eFigure 3a: Kaplan-Meier analysis for overall survival of different dexamethasone dosage patient groups in COHORT A and B**

Kaplan Meier analysis stratified according to the following characteristics: **A)** intake of dexamethasone within the total cohort or COHORT A (n=539) of patients dichotomized into 2 groups: patients that did not receive perioperative dexamethasone (-DEXA in green; median OS was 15.2 months; 95% CI: 9.10-28.9) or those which received dexamethasone during the pre- or postoperative period (+DEXA in red; median OS was 11.2 months; 95% CI: 9.97-13.0) (p=0.36); **B)** intake of dexamethasone within the subcohort COHORT B or “3months and KPS<50excl” (n=483) dichotomized into 2 groups: patients that did not receive perioperative dexamethasone (-DEXA in green; median OS was 16.5 months (95%

CI: 11.2-52.2) or those which received dexamethasone during the pre- or postoperative period (+DEXA in red; median OS was 13.3 months (95% CI: 11.8-15.2) (p=0.32); **C)** intake of dexamethasone within the total cohort (n=539) of patients dichotomized into 2 groups: patients that did receive perioperative dexamethasone below 8 mg daily (< 8mg daily, in green; median OS was 13.0 months (95% CI: 11.17-15.3), or those which received dexamethasone $\geq$ 8 mg daily (during the pre- or postoperative period (+DEXA in red; median OS was 10.5 months (95% CI: 8.43-12.0) (p=0.2); **D)** intake of dexamethasone within the subcohort COHORT B or “3months and KPS<50excl” (n=483) of patients dichotomized into 2 groups: patients that did receive perioperative dexamethasone below 8 mg daily (< 8mg daily, in green; median OS was 14.7 months, 95% CI: 12.9-19.1) or those which received $\geq$ 8 mg dexamethasone daily during the pre- or postoperative period (+DEXA in red; 11.6 months, 95% CI: 10.5-14.9) (p=0.33).

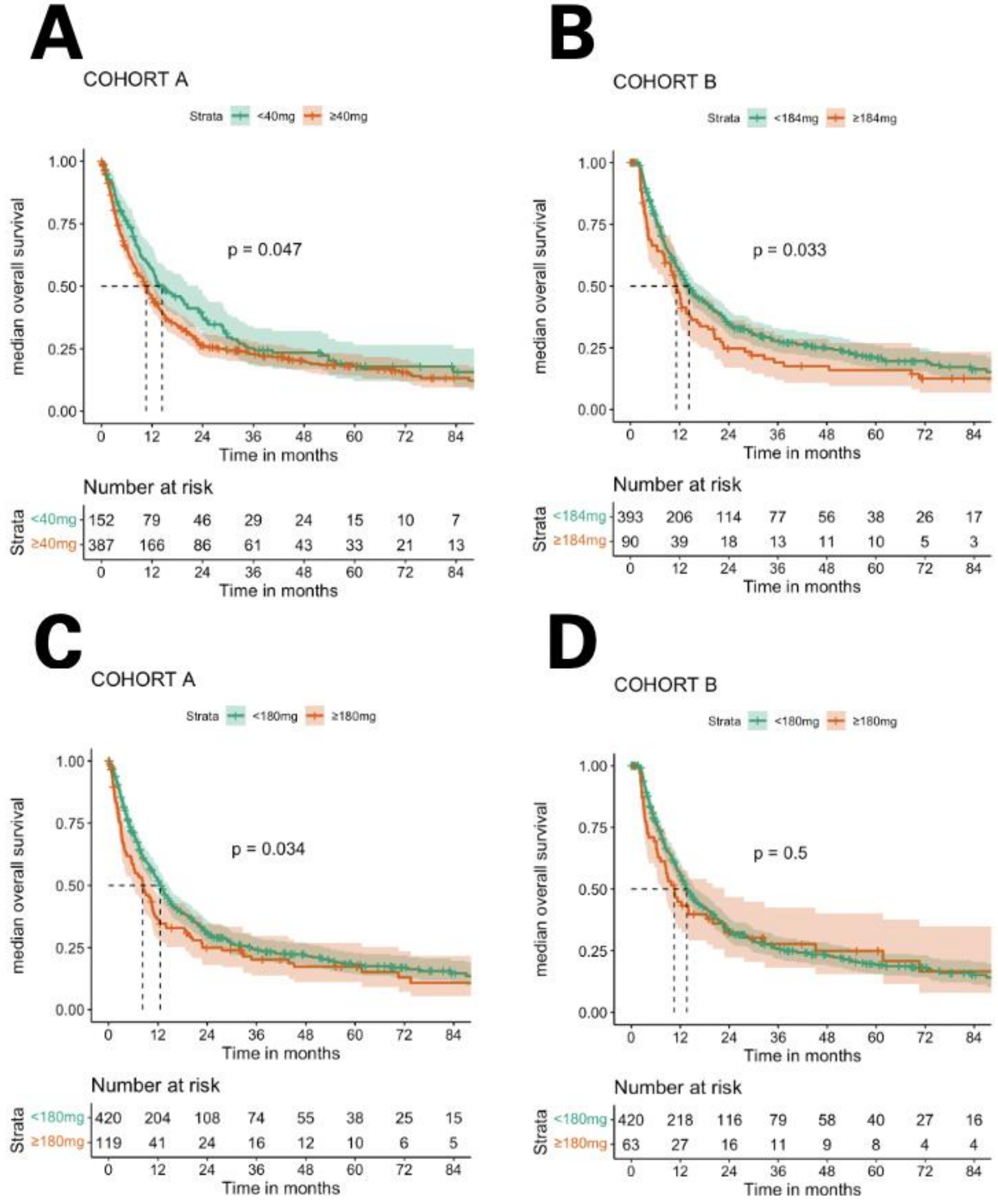


**eFigure 3b: Kaplan-Meier analysis for overall survival of different dexamethasone dosage patient groups in COHORT A and B**

Kaplan Meier analysis stratified according to the following characteristics: **A)** cumulative intake of pre-operative dexamethasone within the total cohort or COHORT A (n=539) or COHORT A dichotomized into 2 groups according to optimal cutpoint of 40 mg: patients with < 40 mg cumulative dexamethasone in the pre-operative period showed a median OS of 14.4 months (95% CI: 11.73-23.0) compared to patients with ≥ 40 mg cumulative dexamethasone in the pre-operative period with an OS of 10.6 months (95% CI: 8.63-12.2) (p=0.047); **B)** In COHORT B patients with < 184 mg cumulative dexamethasone in the pre-operative period showed a median OS of 14.3 months (95% CI: 12.50-17.4) as compared to those patients with ≥ 184 mg cumulative dexamethasone in the pre-operative period with an OS of 11.2 months (95% CI: 8.43-14.4) (p=0.033); **C)** the cutpoint for postoperative dexamethasone dosage was 180 mg which in turn resulted in two subgroups: the patent group with < 180 mg cumulative dexamethasone showed a median OS of 12.6 months (95% CI: 11.03-14.6) vs. those patients with ≥ 180 mg cumulative dexamethasone in the pre-operative period with an OS of 8.33 months (95% CI: 5.77-10.9) (p=0.034); **D)** for the COHORT B optimal cutpoint with 180 mg of post-operative dexamethasone: patients with < 180 mg cumulative dexamethasone (n=420) showed a median OS of 13.6 months (95% CI: 12.30-16.0) vs. patients with ≥ 180 mg cumulative dexamethasone in the post-operative period (n=63) with an OS of 10.5 months (95% CI: 8.23-22.8) (p=0.5).

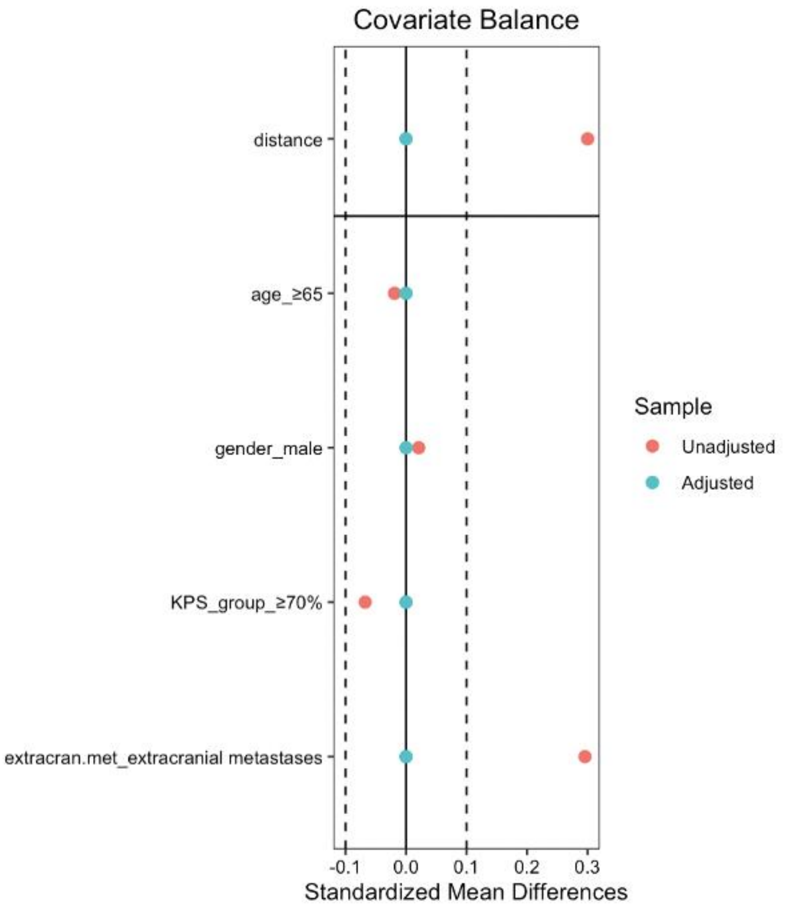


**eFigure 4: Propensity score matching and resulting mean standardized differences**

Distribution of standardized mean differences before (unadjusted) and after (adjusted) propensity score matching (PSM) in COHORT A for covariates considered in the PSM including age, gender, KPS and presence of extracranial metastases.

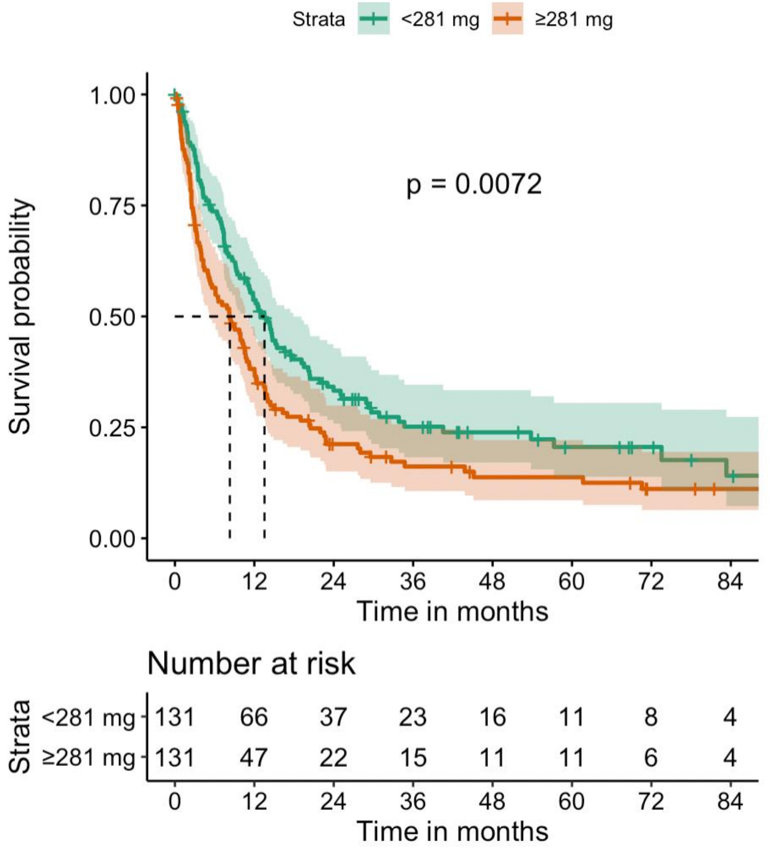


**eFigure 5: Kaplan-Meier survival curve for matched patients after propensity score matching**

Kaplan-Meier curve and associated risk table displaying overall survival of the two cohorts of patients receiving <281 mg perioperative dexamethasone with a median OS of 13.5 months (95% CI: 10.9-17.9) vs. ≥ 281 mg perioperative dexamethasone with a median OS of 8.3 months (95% CI: 5.37-10.9) (p=0.0072).
